# NEWS-2 Score Assessment of Inpatient Referral during the COVID 19 Epidemic

**DOI:** 10.1101/2021.06.10.21254528

**Authors:** Valérie Faure, Marc Souris, Arnaud Wilmet, Franck Baudino, Albert Brizo, Christophe Malhaire, François-Xavier Peccaud, Jean-Paul Gonzalez

## Abstract

**Aim:** To manage patients with suspected coronavirus disease (COVID-19) when they arrive at the hospital emergency department (ED), a clinical severity score is required to quickly identify patients requiring immediate hospital admission and close monitoring. The aim of this study was to evaluate, within the context of the pandemic, the performance of National Early Warning Score 2 (NEWS-2) to anticipate the admission of patients with suspected COVID-19 to a specialised emergency care unit.

**Methods:** This retrospective study was conducted on patients presenting at the COVID-19 entrance of the ED of the Vert-Galant private hospital (Paris, France) during the first national pandemic peak from March 20 to April 20, 2020. All patients completed a questionnaire and clinical data and vital signs were recorded. Statistical analysis and modelling were used to estimate the ability of different scores (NEWS-2, qSOFA, CRB-65) to predict hospital emergency admission and/or early COVID-19 diagnosis.

**Results:** NEWS-2, with a cut off value of 5, predicted hospital admission with 82% sensitivity, 98% specificity and an area under the curve (AUC) of 96%. NEWS-2 was superior to qSOFA and CRB-65 scores for predicting hospital admission of COVID-19 patients. Multilinear or logistic regression analysis of clinical data did not improve this result.

**Conclusion:** NEWS-2 is an excellent score to predict hospital admission of COVID-19 patients.

## Introduction

The COVID-19 pandemic started in early December 2019 in Wuhan, China, and spread worldwide as a severe acute respiratory syndrome caused by the novel coronavirus SARS-CoV-2.^1^ On January 30, 2020, the World Health Organisation (WHO) declared COVID-19 as a Public Health Emergency of International Concern (PHEIC) and designated it as a pandemic on March 11, 2020.^2^ In France, the first case of COVID-19 was identified on January 24, 2020, and “level three” emergency of the COVID epidemic was officially declared by the French authorities on March 14, 2020.

In many countries, the pandemic resulted in a massive influx of patients to hospital emergency departments (ED). The rapid spread of the disease, the increasing number of severe cases requiring hospital admission and the lack of preparation for this unexpected patient influx by the existing healthcare system meant that hospitals had to quickly adapt and modify their strategies for efficient hospital admission.

COVID-19 is associated with a great variety and severity of clinical symptoms; 20% of infected patients who go to hospital will require immediate admission and 5% will be admitted to an intensive care unit (ICU), mainly due to respiratory problems.^3-6^ Although the hospital system in France offers an average of 621 hospital beds per 100 000 inhabitants, in regions of high incidence during the study period most hospitals were overwhelmed by the sudden influx of patients.^7^ In this context, the Vert-Galant Hospital (VGH), a 100-bed private hospital located in Tremblay-en-France, Ile-de-France, 33 km North-East of the centre of Paris, underwent a major organisational change of its ED in order to cope with such an unexpected situation. This reorganisation of the ED encompassed a specific rapid patient pathway that included triage, an initial questionnaire, recording of vital signs with a telemedicine booth (TMB) and a medical examination.

In times of health crisis and a large influx of patients to the ED, clinical scores are often used to rapidly and accurately refer patients. In an evaluation of the changes to be implemented during the epidemic crisis, the use of a standardised and rapid method to assess the severity of COVID-19 patients presenting at the ED was recommended to effectively address the need for admission with rapid hospital triage.^8-10^ Thus, the National Early Warning Score 2 (NEWS-2) that was originally developed as a tool for patient evaluation and monitoring by the Royal College of Physicians in 2012 was identified.^11-13^ All data needed for its calculation are recorded routinely by medical staff. The score aims to improve communication between nurses and physicians, to standardise basic triage practices based on the interpretation of vital signs and, ultimately, to identify patients requiring hospitalisation or intensive care at an early stage.^14-16^ This is reinforced by the instability of vital signs, particularly respiratory, which are a predictive factor for cardiorespiratory aggravation of COVID-19.^17^

The aim of this observational, retrospective, single centre study was to evaluate the performance of NEWS-2 in the disposition of patients with suspected COVID-19 presenting at the ED: either hospital admission (HOS) or discharge home (DH). The secondary aims were to compare NEWS-2 with the qSOFA (quick Sequential Sepsis-related Organ Failure Assessment)^18^ and CRB-65 (Mental Confusion, Respiratory rate, Blood pressure, Age)^19^ scores, and to evaluate the concordance between NEWS-2 and pulmonary computed tomography (CT) scans as a marker of disease severity. The final aim was to determine whether NEWS-2 can be considered as an aid to COVID-19 diagnosis.

## Methods

### Study design and population

All patients included in this study were adults (≥18-years-old) and presented for triage at the ED of the COVID sector of VGH between March 19 and April 29, 2020. The exclusion criteria included: more than one presentation during the recruitment period; hospital admission despite the lack of clinical severity due to social reasons or if discharging the patient back home was not possible; protected patient including: adult under guardianship or other legal protection, deprived of liberty by judicial or administrative decree; pregnancy or breastfeeding; hospital admission without consent; and missing data preventing calculation of NEWS-2. All patients presenting at the ED with at least one of the respiratory and gastrointestinal signs and symptoms of COVID-19, as described in the literature, were included in the study. They represent 41% of the total ED population during the study period.^20^

### Ethics and data protection

All patients were informed of the data collection by a specific poster in the waiting room and gave their informed consent prior to the recording of vital signs by the TMB. All patients received a letter informing them of the study and gave their informed consent, according to MR004 methodology.

The project, (referenced under the number COS-RGDS-2021-05-001-FAURE-V) has been submitted to the members of the Scientific Committee of the GCS Ramsay Santé for Education and Research. The committee has reviewed the following documents: application form for ethical advice on a project research and the synopsis of the study. This study has received a favorable opinion on 05/04/2021 (Numéro IORG de Ramsay santé Recherche & Enseignement: IORG0009085; Numéro IRB du Conseil d’Orientation Scientifique: IRB00010835).

### Data collection

Demographic and clinical data, vital signs and details of patient disposition were collected retrospectively from the electronic medical records. During the day, a Class 2 certified TMB was used, allowing the autonomous measurement and automatic recording of vital signs (pulse rate, oxygenation, systolic pressure (SBP), diastolic pressure (DBP), pulse pressure, mean arterial pressure (MAP), temperature and respiratory rate). The TMB equipment (ConsultStation®) was generously provided by H4D Inc., Paris, France. The data collected by the TMB were copied into the electronic record.

Data for mobile patients were collected by the TMB, while data for bed-bound patients were collected by the nursing staff. The clinical signs recorded included: changes in consciousness; breathlessness; altered general condition (defined as a clinical condition where the vital signs were not significantly altered, but other clinical symptoms such as intense asthenia, vomiting and anorexia (difficulty eating and drinking during the previous 24 h) pointed to hospital admission. By convention, when a figure for the respiratory rate was not available and the physician reported the respiration of the patient as “normal” or “eupnoeic” on the report, it was quantified as 16 breaths min^-1^. All data, including that from the medical and paramedical examination and data collected by the TMB, were analysed anonymously.

### Scores and complementary examinations

Three clinical scores were calculated and compared: NEWS-2, qSOFA and CRB-65.

NEWS-2 is based on the measurement of six physiological parameters: respiratory rate, oxygen saturation, SBP, pulse rate, level of consciousness and body temperature. Each parameter is allocated a sub-score as it is measured, with the magnitude reflecting how much the parameter varies from the norm. The sub-scores are then added together as indicated in the latest version of NEWS-2.^13^ NEWS-2 score can range from 0 to 20, with a score of ≥7 indicating the need for a rapid clinical assessment and potential transfer of the patient to an ICU.

The qSOFA score is used to identify a possible infectious syndrome. Total score ranges from 0 to 3 with one point each allocated for low blood pressure (SBP ≤100 mmHg), high respiratory rate (≥22 breaths min^-1^) and altered consciousness (Glasgow coma scale <15).

Finally, the CBR-65 score is used to assess the severity of community-acquired pneumonia and to determine whether a patient requires inpatient treatment or not. Total CRB-65 score ranges from 0 to 4, with one point each being allocated for: mental confusion, respiratory rate ≥30 breaths min^-1^, SBP <90 mmHg or DBP ≤60 mmHg, and age ≥65 years.

Other tests included RT-PCR for the diagnosis of COVID-19 and a pulmonary CT scan interpreted according to the Society for Thoracic Imaging (STI).^21^

### Evaluation criteria

The primary evaluation criterion was the concordance between NEWS-2 and real disposition of the patient from the ED: HOS or DH. The secondary objectives were to compare NEWS-2 with the qSOFA and CRB-65 scores for patient disposition and to determine whether NEWS-2 can be considered as an aid to COVID-19 diagnosis.

### Statistical analysis

The ability of NEWS-2 to predict HOS or DH was first assessed by descriptive and exploratory analyses. Sensitivity/specificity analysis and receiver operating characteristic (ROC) curves were used to explore NEWS-2 performance and to detect the best cut-off value for NEWS-2 as a predictor of patient disposition and COVID-19 pre-diagnosis severity. To avoid problems with multiple testing, *P* values were adjusted using Bonferroni correction to keep the false discovery rate (α-risk) at 5%. Discriminant analysis or statistical models based on multilinear or logistic regression were used to determine whether adding weights to sub-scores or adding new parameters (e.g., comorbidities, anorexia, age, general condition of the patient) could improve the performance of NEWS-2 for predicting hospital disposition or severity for COVID-19 positive patients.

All statistical calculations were performed with XLSTAT software (version 2020.5.1, Addinsoft) and SavGIS software (version 9.07.015, Institut de Recherche pour le Développement, IRD).

## Results

### Study population

A total of 853 patients, chosen after initial triage, were included in the study population. Thirty-nine declined to take part in the study, thus 814 patients were included in the final analysis. Mean age was 46.8 years, with a M/F sex ratio of 0.5. Almost two-thirds (60%) of the patients visited the TMB to record their vital signs. The comorbidities of the patients included, arterial hypertension (HA; 18.9% of all patients), cardiovascular disease (Cardio; 6.8%), diabetes (10.0%) and chronic obstructive pulmonary disease (COPD; 1.6%). Nearly one-quarter (23.7%) of patients had at least one comorbidity (14.2% had a single comorbidity, 6.8% had two comorbidities, 2.6% had three comorbidities and 0.2% had four comorbidities).

### COVID-19 pre-diagnosis

Within the context of the epidemic, COVID-19 was diagnosed clinically by the emergency doctor in two-thirds (n=543, 67%) of the 814 patients. A total of 428/814 patients were tested by RT-PCR, a pulmonary CT scan, or both. A positive diagnosis of COVID-19 was made in 246/428 (57.5%) tested patients. The demographic and clinical data for the study participants are summarised in Table 1.

**Table 1.** Demographic and clinical data for the study population (TMB: telemedicine booth; COPD: chronic obstructive pulmonary disease; HA: arterial hypertension; Cardio: cardiovascular disease; SBP: systolic blood pressure; DBP: diastolic blood pressure; MAP: mean arterial blood pressure; BMI: body mass index).

|  | Percentage or Mean | Missing data |
| --- | --- | --- |
| Age (years) (Mean) | 46.81 | 0 |
| Sex (male) | 50.0% | 0 |
| Data recorded by TMB | 60.0% | 20 |
| **Comorbidities** |  |  |
| COPD | 1.6% | 4 |
| HA | 18.9% | 0 |
| Cardio | 6.8% | 0 |
| Diabetes | 10.0% | 26 |
| **No. of comorbidities** |  |  |
| 0 | 76.3% | 30 |
| 1 | 14.2% | 30 |
| 2 | 6.8% | 30 |
| 3 | 2.6% | 30 |
| 4 | 0.2% | 30 |
| **Vital signs** |  |  |
| Pulse rate (beats/min) (Mean) | 90.93 | 8 |
| SpO2 (%) (Mean) | 96.64 | 4 |
| SBP (mmHg) (Mean) | 136.41 | 9 |
| DBP (mmHg) (Mean) | 84.43 | 9 |
| MAP (mmHg) (Mean) | 101.76 | 9 |
| Pulse pressure (mmHg) (Mean) | 51.40 | 0 |
| Temperature, °C (Mean) | 37.21 | 7 |
| Respiratory rate (breaths/min) (Mean) | 19.23 | 188 |
| BMI (kg/m2) (Mean) | 27.09 | 347 |
| **Symptoms/signs** |  |  |
| Vomiting | 20.2% | 507 |
| Anorexia | 22.5% | 467 |
| Altered general state | 15.2% | 129 |
| **Diagnosis** |  |  |
| COVID-19 | 61.0% | 9 |
| Reliability (1-4) | 2 | 9 |
| **Orientation** |  |  |
| Hospital admission | 14.0% | 0 |
| Return home | 86.0% | 0 |
| Scores |  |  |
| NEWS-2 (Mean) | 2.36 | 194 |
| qSOFA (Mean) | 0.26 | 191 |
| CRB65 (Mean) | 0.32 | 191 |

NEWS-2 score ranged from 0 to 13 with a mean of 2.36, a bimodal distribution and a natural cut-off between 4 and 5 (Fig. 1).

**Figure 1.**
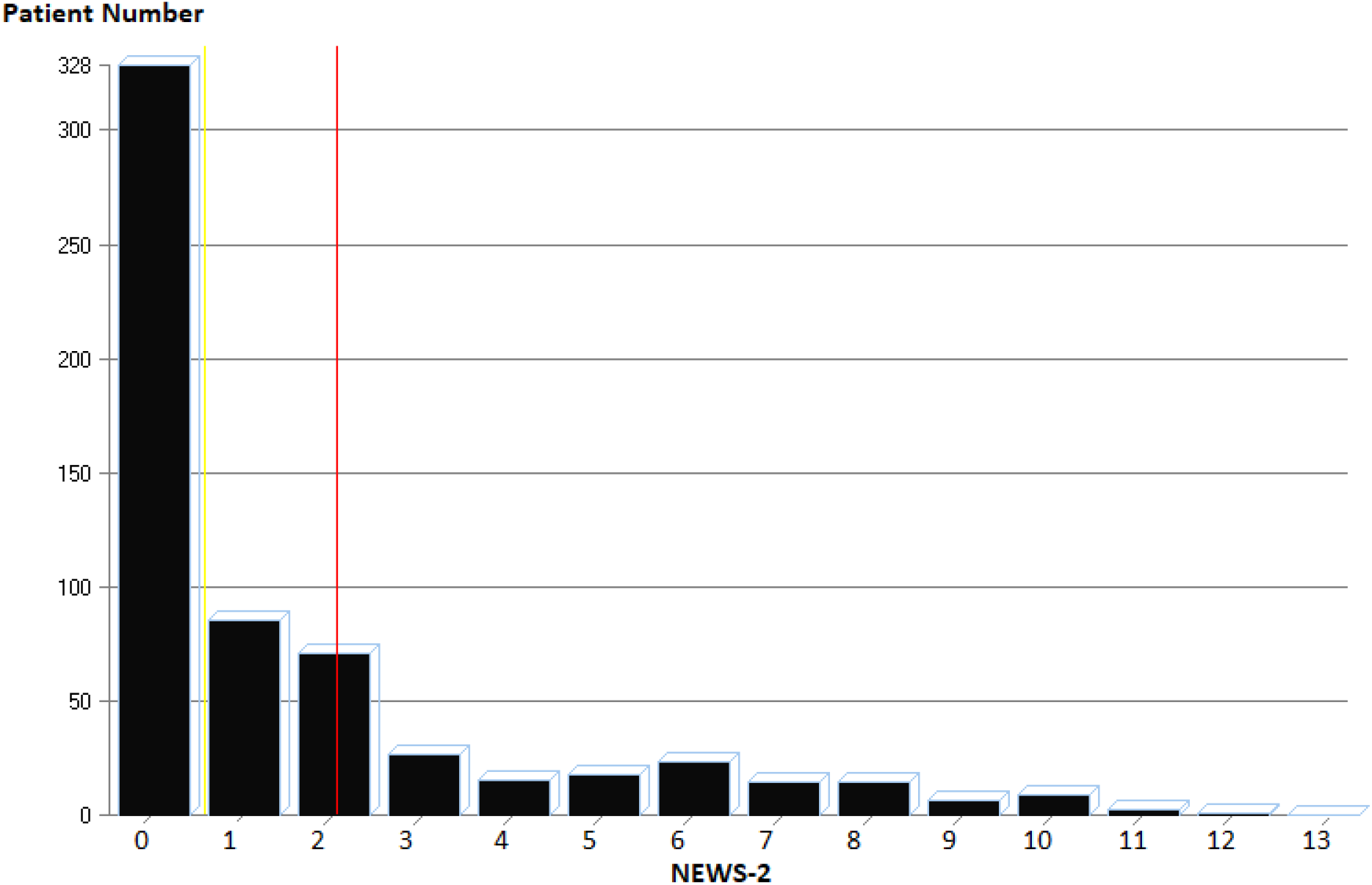
Distribution of NEWS-2 values in the study population. Mean value of 2.36 (red line).

### Patient disposition and scores

After the initial triage, a second triage was carried out in the ED, assessing the vital signs and general condition of the patient, to determine the disposition of the patient: HOS or DH. The patient disposition chosen by the ED team served as the reference to evaluate the three scores (NEWS-2, qSOFA, CRB-65). Among the 814 patients, 114 (14.0%) were HOS and 700 (86.0%) were DH. Most of the variables examined differed significantly between the two groups (Table 2). The greatest difference between the two groups was “altered general condition” (p-value <10^−6^). Age and sex were also significantly different between the two groups but these variables are often considered as confounding factors (age nevertheless remains a risk factor for severity and death in the context of COVID-19 even after removing the comorbidities associated with it). Odds ratios were also calculated after adjusting for sex and age (>65-years).^22,23^

**Table 2.**
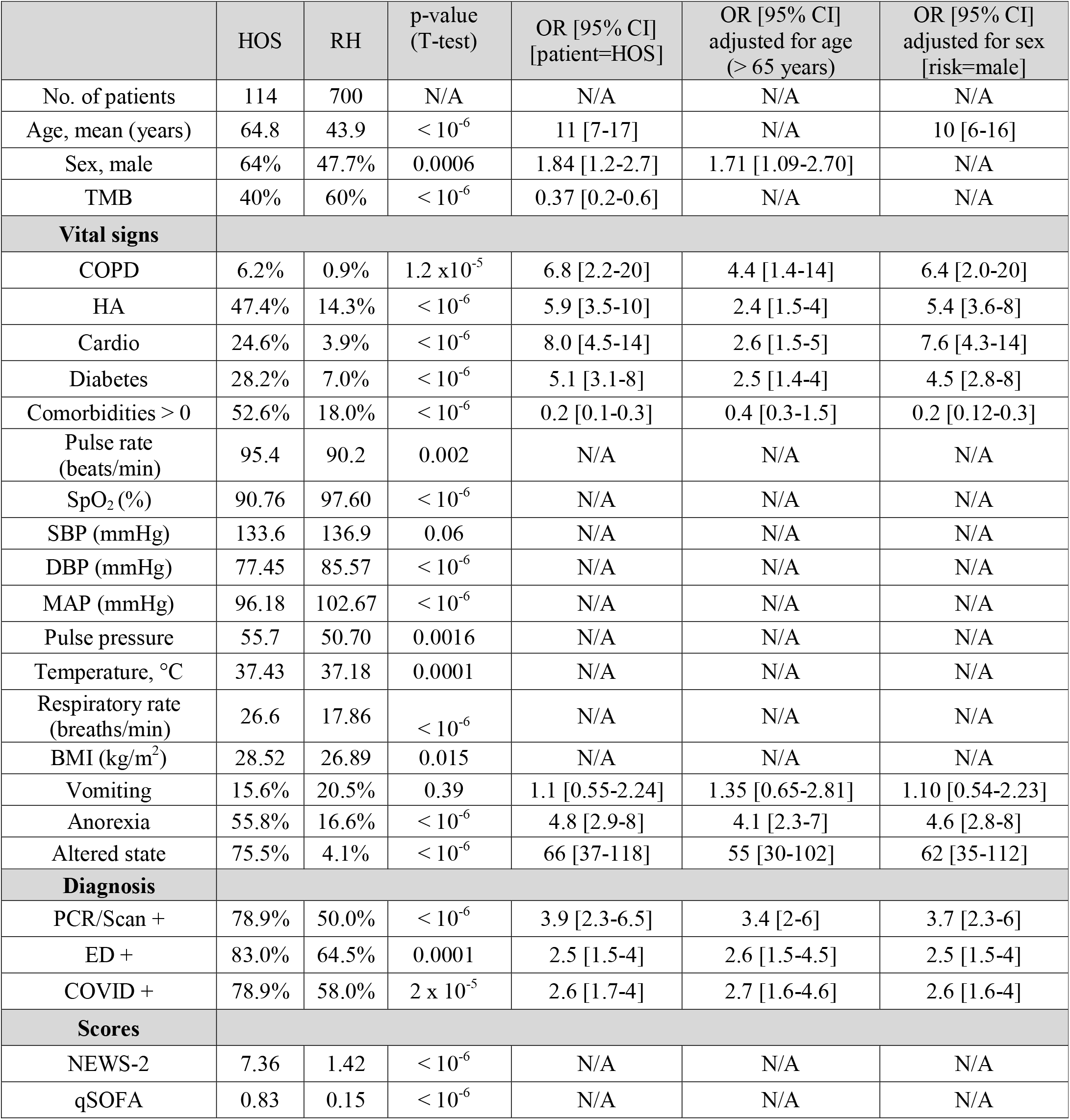
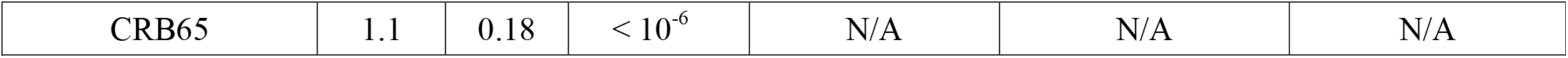
Comparison of patients admitted to hospital (HOS) and those who returned home (RH) (CI: Confidence Interval; TMB: Telemedicine Booth; COPD: chronic obstructive pulmonary disease; HA: arterial hypertension; Cardio: cardiovascular disease; SBP: systolic blood pressure; DBP: diastolic blood pressure; MAP: mean arterial blood pressure; BMI: body mass index; ED: emergency department. N/A: not applicable)

|  | HOS | RH | p-value<br>(T-test) | OR [95% CI]<br>[patient=HOS] | OR [95% CI]<br>adjusted for age<br>(> 65 years) | OR [95% CI]<br>adjusted for sex<br>[risk=male] |
| --- | --- | --- | --- | --- | --- | --- |
| No. of patients | 114 | 700 | N/A | N/A | N/A | N/A |
| Age, mean (years) | 64.8 | 43.9 | $< 10^{-6}$ | 11 [7-17] | N/A | 10 [6-16] |
| Sex, male | 64% | 47.7% | 0.0006 | 1.84 [1.2-2.7] | 1.71 [1.09-2.70] | N/A |
| TMB | 40% | 60% | $< 10^{-6}$ | 0.37 [0.2-0.6] | N/A | N/A |
| **Vital signs** |  |  |  |  |  |  |
| COPD | 6.2% | 0.9% | $1.2 \times 10^{-5}$ | 6.8 [2.2-20] | 4.4 [1.4-14] | 6.4 [2.0-20] |
| HA | 47.4% | 14.3% | $< 10^{-6}$ | 5.9 [3.5-10] | 2.4 [1.5-4] | 5.4 [3.6-8] |
| Cardio | 24.6% | 3.9% | $< 10^{-6}$ | 8.0 [4.5-14] | 2.6 [1.5-5] | 7.6 [4.3-14] |
| Diabetes | 28.2% | 7.0% | $< 10^{-6}$ | 5.1 [3.1-8] | 2.5 [1.4-4] | 4.5 [2.8-8] |
| Comorbidities > 0 | 52.6% | 18.0% | $< 10^{-6}$ | 0.2 [0.1-0.3] | 0.4 [0.3-1.5] | 0.2 [0.12-0.3] |
| Pulse rate<br>(beats/min) | 95.4 | 90.2 | 0.002 | N/A | N/A | N/A |
| SpO<sub>2</sub> (%) | 90.76 | 97.60 | $< 10^{-6}$ | N/A | N/A | N/A |
| SBP (mmHg) | 133.6 | 136.9 | 0.06 | N/A | N/A | N/A |
| DBP (mmHg) | 77.45 | 85.57 | $< 10^{-6}$ | N/A | N/A | N/A |
| MAP (mmHg) | 96.18 | 102.67 | $< 10^{-6}$ | N/A | N/A | N/A |
| Pulse pressure | 55.7 | 50.70 | 0.0016 | N/A | N/A | N/A |
| Temperature, °C | 37.43 | 37.18 | 0.0001 | N/A | N/A | N/A |
| Respiratory rate<br>(breaths/min) | 26.6 | 17.86 | $< 10^{-6}$ | N/A | N/A | N/A |
| BMI (kg/m<sup>2</sup>) | 28.52 | 26.89 | 0.015 | N/A | N/A | N/A |
| Vomiting | 15.6% | 20.5% | 0.39 | 1.1 [0.55-2.24] | 1.35 [0.65-2.81] | 1.10 [0.54-2.23] |
| Anorexia | 55.8% | 16.6% | $< 10^{-6}$ | 4.8 [2.9-8] | 4.1 [2.3-7] | 4.6 [2.8-8] |
| Altered state | 75.5% | 4.1% | $< 10^{-6}$ | 66 [37-118] | 55 [30-102] | 62 [35-112] |
| **Diagnosis** |  |  |  |  |  |  |
| PCR/Scan + | 78.9% | 50.0% | $< 10^{-6}$ | 3.9 [2.3-6.5] | 3.4 [2-6] | 3.7 [2.3-6] |
| ED + | 83.0% | 64.5% | 0.0001 | 2.5 [1.5-4] | 2.6 [1.5-4.5] | 2.5 [1.5-4] |
| COVID + | 78.9% | 58.0% | $2 \times 10^{-5}$ | 2.6 [1.7-4] | 2.7 [1.6-4.6] | 2.6 [1.6-4] |
| **Scores** |  |  |  |  |  |  |
| NEWS-2 | 7.36 | 1.42 | $< 10^{-6}$ | N/A | N/A | N/A |
| qSOFA | 0.83 | 0.15 | $< 10^{-6}$ | N/A | N/A | N/A |
| CRB65 | 1.1 | 0.18 | $< 10^{-6}$ | N/A | N/A | N/A |

To evaluate NEWS-2 as an aid for the disposition of COVID-19 patients, the discriminatory power of the score for hospital admission was verified, evaluating the best cut-off value and analysing the patients who were misclassified by the score (Fig. 2). NEWS-2 with its optimum cut-off was then considered as a test for hospital admission, to measure its sensitivity and specificity. Among the 620 patients for whom we could calculate NEWS-2, 98 (15.8%) were HOS and 522 (84.2%) were DH. Even though it did not take into account comorbidities or ‘altered general condition’, NEWS-2 was significantly different between the two groups (mean 7.36 for HOS vs. 1.42 for DH; p-value <10^−6^). The sensitivity, specificity and predictive values for the different cut-off values of NEWS-2 are shown in Table 3. The cut-off value of 5 offered the best sensitivity (82%) and specificity (98%). This value minimised the number of patients that were misclassified (n=28) (17 false negatives: NEWS-2 score ≤5 but patients HOS; 11 false positives: NEWS-2 score >5 but patients DH).

**Table 3.** Sensitivity, specificity, and predictive values for the different cut-off values for NEWS-2 in relation to Hospital admission (TP: true positive; FP: false positive; TN: true negative; FN: false negative; PVP: predictive value positive; PVN: predictive value negative; LR+: positive likelihood ratio = sensitivity / (1 − specificity); LR-: negative likelihood ratio = (1 − sensitivity) / (specificity)).

| NEWS-2<br>Cut-off | TP | FP | TN | FN | Sensitivity | Specificity | PVP | PVN | LR+ | LR- |
| --- | --- | --- | --- | --- | --- | --- | --- | --- | --- | --- |
| 3 | 88 | 47 | 475 | 10 | 0.898 | 0.910 | 0.65 | 0.99 | 9.97 | 0.11 |
| 4 | 84 | 24 | 498 | 14 | 0.857 | 0.954 | 0.78 | 0.97 | 18.6 | 0.15 |
| 5 | 81 | 11 | 511 | 17 | 0.827 | 0.979 | 0.88 | 0.96 | 39.2 | 0.17 |
| 6 | 68 | 6 | 516 | 30 | 0.694 | 0.989 | 0.92 | 0.95 | 60.42 | 0.31 |
| 7 | 46 | 4 | 518 | 52 | 0.469 | 0.992 | 0.92 | 0.91 | 61.25 | 0.53 |
| 8 | 32 | 3 | 519 | 66 | 0.327 | 0.994 | 0.91 | 0.89 | 56.82 | 0.68 |

**Figure 2.**
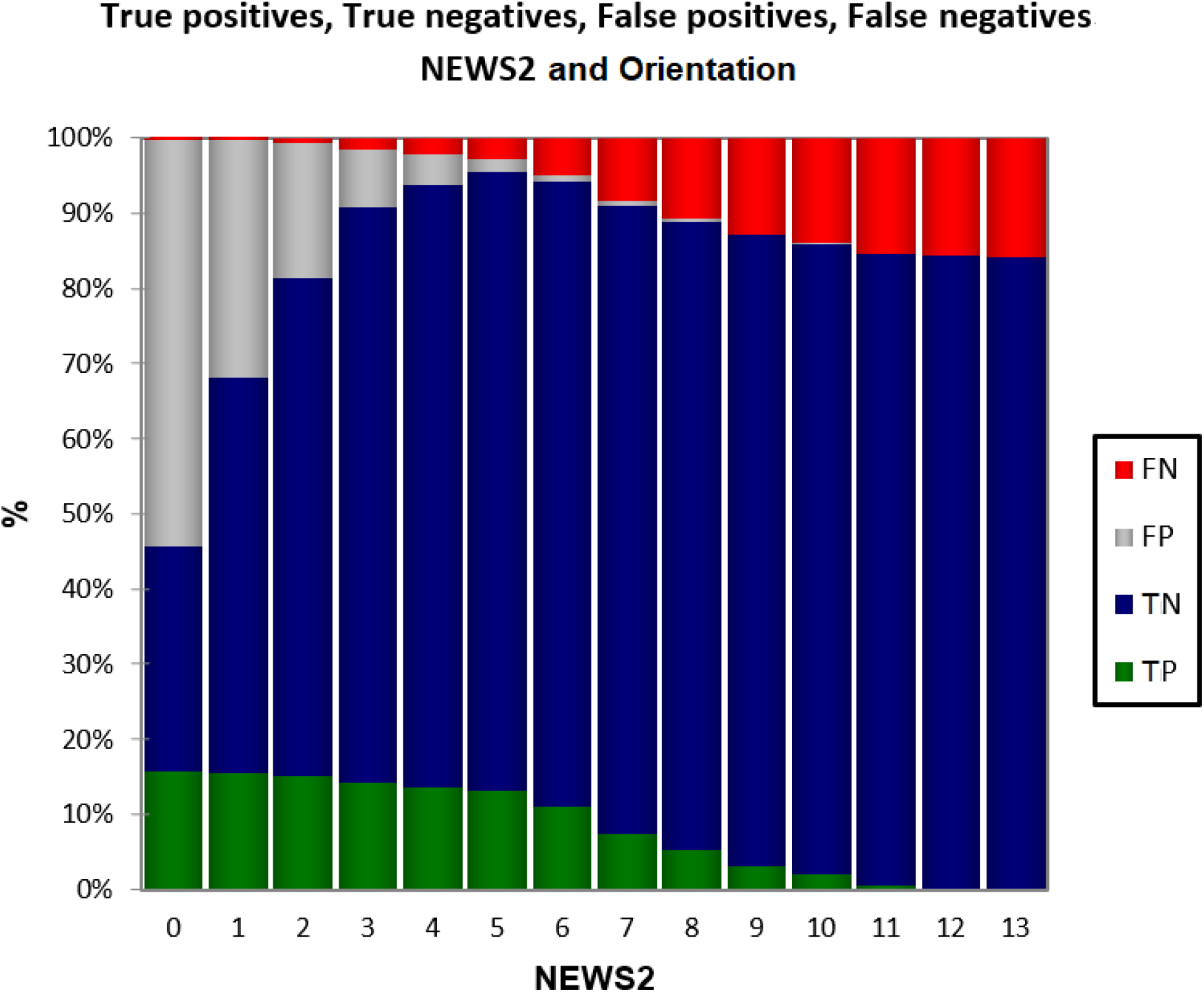
Different NEWS-2 cut-off values for hospital admission of COVID-19 patients.

ROC curves of the three scores showed that NEWS-2 was the most predictive for hospital admission (AUC = 0.96) (Fig. 3).

**Figure 3.**
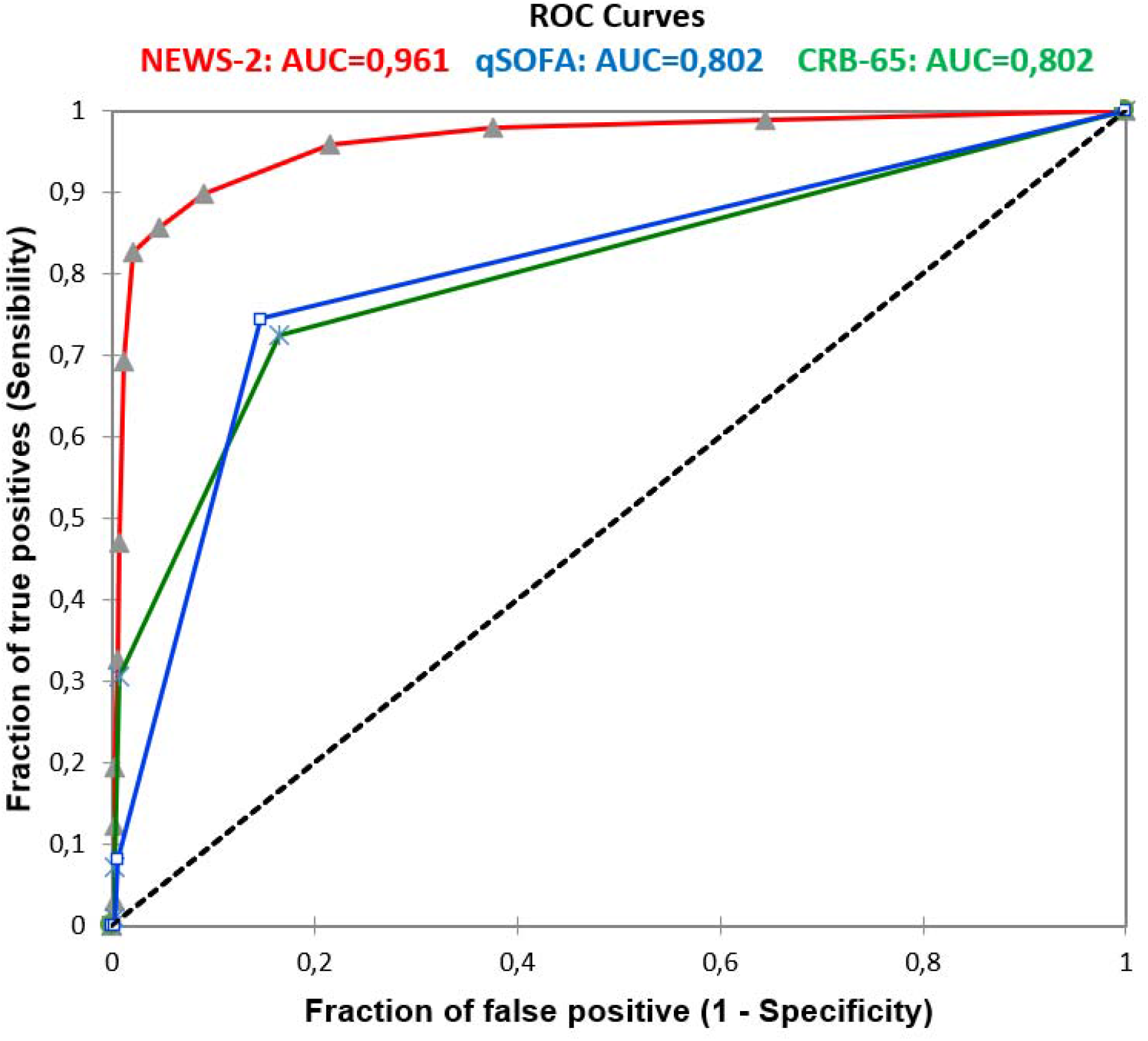
Comparison of sensitivity and specificity of NEWS-2, qSOFA and CRB-65 scores as a test for hospital admission of COVID-19 patients.

In order to understand why 17 patients with NEWS-2 ≤5 were admitted to hospital by the ED, we investigated the differences between the HOS group with NEWS-2 = 4 or 5 (false negatives) and DH with NEWS-2 = 4 or 5 (true negatives). Age, anorexia and ‘altered general condition’ were the most discriminating factors for HOS that were not directly included in NEWS-2. In order to understand why 11 patients with a score >5 were not admitted to hospital, we investigated the differences between the DH group with NEWS-2 = 6 or 7 (false negatives) and the HOS group with NEWS-2 = 6 or 7 (true positives). Altogether, age and sex were significant factors for DH (in the absence of comorbidities, principally HA and diabetes). The presence of vomiting was not a criterion for HOS (Table 4).

**Table 4.** Differences in variables between true and false negatives, NEWS-2 score of 4 or 5 and, between true and false positives, NEWS-2 score of 6 or 7 (COPD: chronic obstructive pulmonary disease; HA: arterial hypertension; Cardio: cardiovascular disease; SBP: systolic blood pressure; DBP: diastolic blood pressure; MAP: mean arterial blood pressure; BMI: body mass index; ED: emergency department; TP: true positive; FP: false positive; TN: true negative; FN: false negative; RH returned home; HOS: hospital admission).

|  | NEWS-2 score4<br>or 5 and HOS<br>(FN) | NEWS-2<br>score 4 or<br>5 and RH<br>(TN) | p-value | NEWS-2<br>score 6 or 7<br>and RH (FP) | NEWS-2 score6<br>or 7 and HOS<br>(TP) | p-value |
| --- | --- | --- | --- | --- | --- | --- |
| No, of patients | 7 | 36 |  | 7 | 35 |  |
| Age (years), mean | 59.4 | 48.8 | 0.05 | 47.7 | 65.5 | 0.004 |
| Sex (male), (%) | 57% | 53% | 0.4 | 43% | 71% | 0.08 |
| **Comorbidities** |  |  |  |  |  |  |
| COPD | 0% | 0% | - | 0% | 5.7% | 0.26 |
| HA | 57% | 19% | 0.02 | 14.3% | 57% | 0.02 |
| Cardio | 0% | 0% | - | 14.3% | 31.5% | 0.18 |
| Diabetes | 0% | 6% | 0.26 | 0% | 34% | 0.03 |
| Comorbidities | 0.57 | 0.24 | 0.07 | 0.29 | 1.29 | 0.02 |
| **Vital signs** |  |  |  |  |  |  |
| Pulse rate<br>(beats/min) | 87.28 | 101.94 | 0.02 | 96 | 90.77 | 0.25 |
| SpO<sub>2</sub> (%) | 94.43 | 96.72 | 0.02 | 95.85 | 92.88 | 0.02 |
| SBP (mmHg) | 132.71 | 133.58 | 0.46 | 144 | 131 | 0.16 |
| DBP (mmHg) | 81.29 | 86.08 | 0.23 | 81.9 | 75.7 | 0.17 |
| MAP (mmHg) | 98.43 | 101.91 | 0.31 | 102.54 | 94.41 | 0.15 |
| Pulse pressure<br>(mmHg) | 51.43 | 47.5 | 0.24 | 62 | 56 | 0.26 |
| Temperature, °C | 37.31 | 37.25 | 0.43 | 37.8 | 37.5 | 0.23 |
| Respiratory rate<br>(breaths/min) | 19.85 | 24.77 | 0.02 | 26 | 23.2 | 0.07 |
| BMI | 29.32 | 29.19 | 0.48 | 24.6 | 29.8 | 0.08 |
| Vomiting | 0% | 14% | 0.20 | 66% | 10% | 0.02 |
| Anorexia | 40% | 0% | 0.002 | 100% | 50% | 0.06 |
| Altered state | 57% | 3% | 10<sup>-5</sup> | 16% | 70% | 0.006 |
| **COVID**<br>**Diagnosis** |  |  |  |  |  |  |
| PCR/Scan+ | 71.4% | 72% | 0.47 | 75% | 89% | 0.22 |
| ED+ | 85% | 75% | 0.30 | 75% | 91% | 0.10 |
| COVID+ | 71.4% | 72.2% | 0.43 | 62% | 89% | 0.03 |

| Scores |  |  |  |  |  |  |
| --- | --- | --- | --- | --- | --- | --- |
| qSOFA | 0.28 | 0.81 | 0.002 | 1.1 | 0.74 | 0.03 |
| CRB65 | 0.43 | 0.47 | 0.42 | 0.29 | 0.8 | 0.04 |

When the different scores were compared, qSOFA did not give any benefit when compared to NEWS-2. Likewise, CRB-65 did not result in any improvement in the false negatives, but clearly differentiated the false positives and true positives for NEWS-2 (6 and 7) values (close to the best cut-off value of 5 for NEWS-2). For these two scores, the reliability of COVID-19 diagnosis seemed to play a role since the diagnosis (confirmed by PCR or CT scan) supported hospital admission of the patient when there was a contradiction between NEWS-2 and disposition (Table 4).

### COVID-19 diagnosis

After the initial triage, almost all patients received a clinical diagnosis from the emergency doctors (799/814 patients) and some were tested by RT-PCR (222/814) or pulmonary CT scan (370/814). A COVID-19 diagnosis was therefore established: positive = RT-PCR positive, or if not, CT scan positive (confirmed diagnosis); negative = CT scan negative (confirmed diagnosis), and if no scan performed, diagnosis based on clinical signs (clinical diagnosis). We therefore obtained 491 positive and 314 negative patients. Few vital signs were discriminatory for COVID-19 diagnosis (apart from SpO_2_, pulse rate and ‘altered general condition’) (Table 5). Nearly 16% of positive clinical diagnoses were contradicted by a negative CT scan.

**Table 5.** Comparison of clinical characteristics between the positive and negative groups for COVID-19 diagnostic (CI: confidence interval; TMB = telemedicine booth; COPD: chronic obstructive pulmonary disease; HA: arterial hypertension; Cardio: cardiovascular disease; SBP: systolic blood pressure; DBP: diastolic blood pressure; MAP: mean arterial blood pressure; BMI: body mass index).

|  | Positive | Negative | p-value (Student's T-test) | OR [95% CI] |
| --- | --- | --- | --- | --- |
| No. of patients | 491 | 314 |  |  |
| Age (years), mean | 47.46 | 45.60 | 0.06 | N/A |
| Sex, male | 53% | 46% | 0.024 | 1.29 [0.98-1.70] |
| TMB | 60% | 61% | 0.4 | 0.94 [0.71-1.26] |
| COPD | 0.8% | 2.8% | 0.01 | 0.25 [0.08-0.81] |
| HA | 18.7% | 19.4% | 0.40 | 0.96 [0.67-1.37] |
| Cardio | 4.9% | 9.2% | 0.007 | 0.46 [0.27-0.80] |
| Diabetes | 11% | 7.6% | 0.04 | 1.6 [0.98-2.61] |
| Comorbidity | 0.36 | 0.38 | 0.35 | 0.93 [0.67-1.30] |
| **Vital signs** |  |  |  |  |
| Pulse rate (beats/min) | 91.08 | 91.0<br>1 | 0.45 | N/A |
| SpO<sub>2</sub> (%) | 96.18 | 97.3<br>1 | 10<sup>-5</sup> | N/A |
| SBP (mmHg) | 135.6 | 137.<br>6 | 0.08 | N/A |
| DBP (mmHg) | 83.97 | 85.2<br>1 | 0.11 | N/A |
| MAP (mmHg) | 101 | 102.<br>7 | 0.08 | N/A |
| Pulse pressure (mmHg) | 51.07 | 51.7<br>7 | 0.28 | N/A |
| Temperature °C | 37.26 | 37.1<br>4 | 0.006 | N/A |
| Respiratory rate (breaths/min) | 19.91 | 17.9<br>2 | 10<sup>-5</sup> | N/A |
| BMI | 27.34 | 26.7<br>2 | 0.12 | N/A |
| Vomiting | 18.9<br>% | 21.7<br>% | 0.3 | 0.73 [0.44-1.21] |
| Anorexia | 27.22<br>% | 15.8<br>6% | 0.006 | 1.77 [1.05-2.93] |
| Altered state | 18.9<br>% | 9.2<br>% | 0.0003 | 2.40 [1.49-3.87] |
| **Scores** |  |  |  |  |
| NEWS-2 | 2.71 | 1.72 | 10<sup>-5</sup> |  |
| qSOFA | 0.31 | 0.17 | 0.0006 |  |
| CRB65 | 0.35 | 0.28 | 0.09 |  |

### COVID-19 diagnosis and NEWS-2

Although NEWS-2 was higher in COVID+ patients than in COVID-patients (mean 2.71 vs. 1.72, respectively), it was not a good predictive test for diagnosis, whatever the chosen cut-off value (Fig. 4). For example, out of 414 patients with NEWS-2 ≤2, 248 were diagnosed as COVID+ (60%) and of 528 patients with NEWS-2 ≤5, 327 were diagnosed as COVID+ (62%). The ROC curve of sensitivity vs. specificity of NEWS-2 had an AUC of 0.607 (Fig. 5).

**Figure 4.**
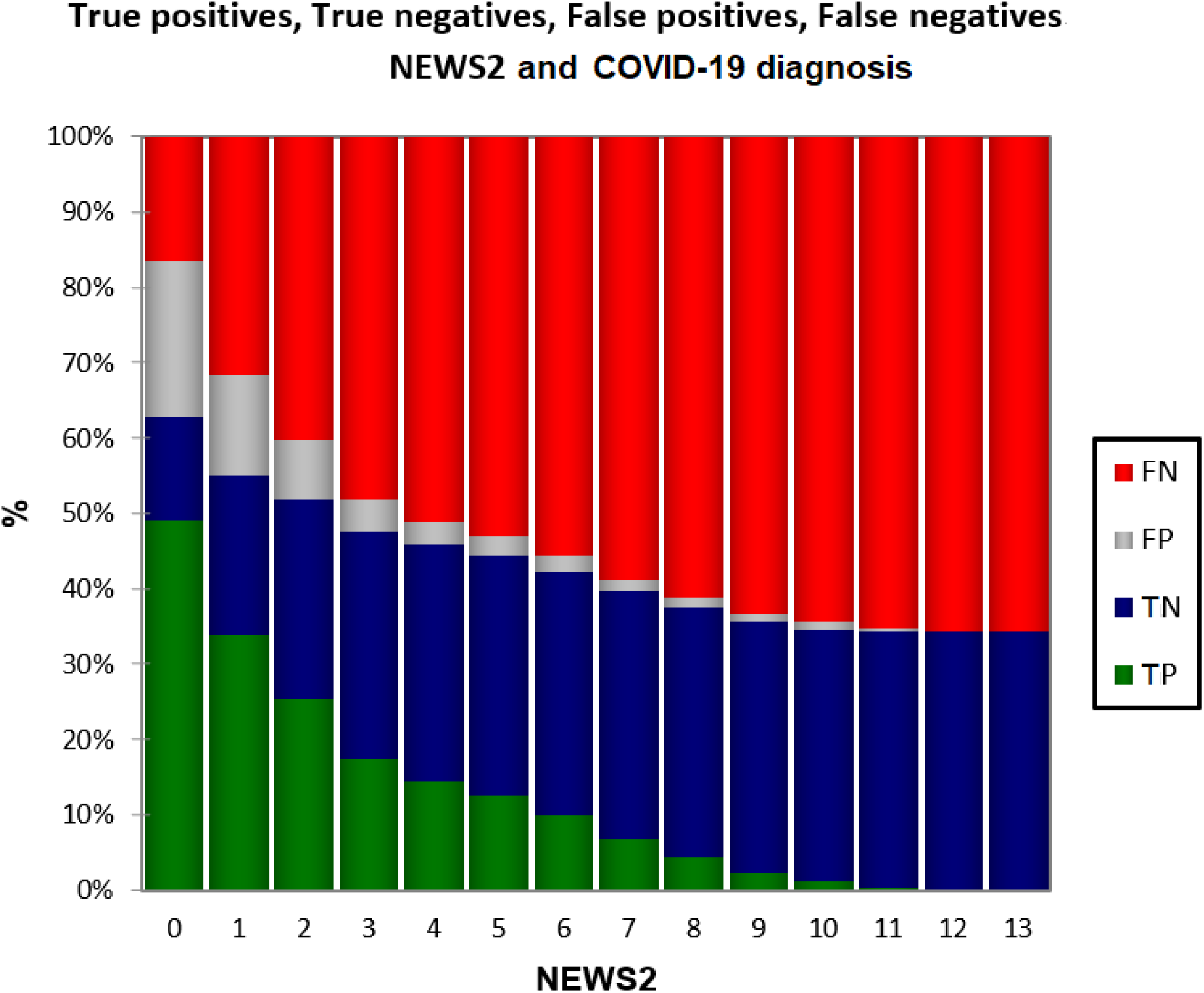
True and false COVID-19 diagnoses in relation to NEWS-2 score.

**Figure 5.**
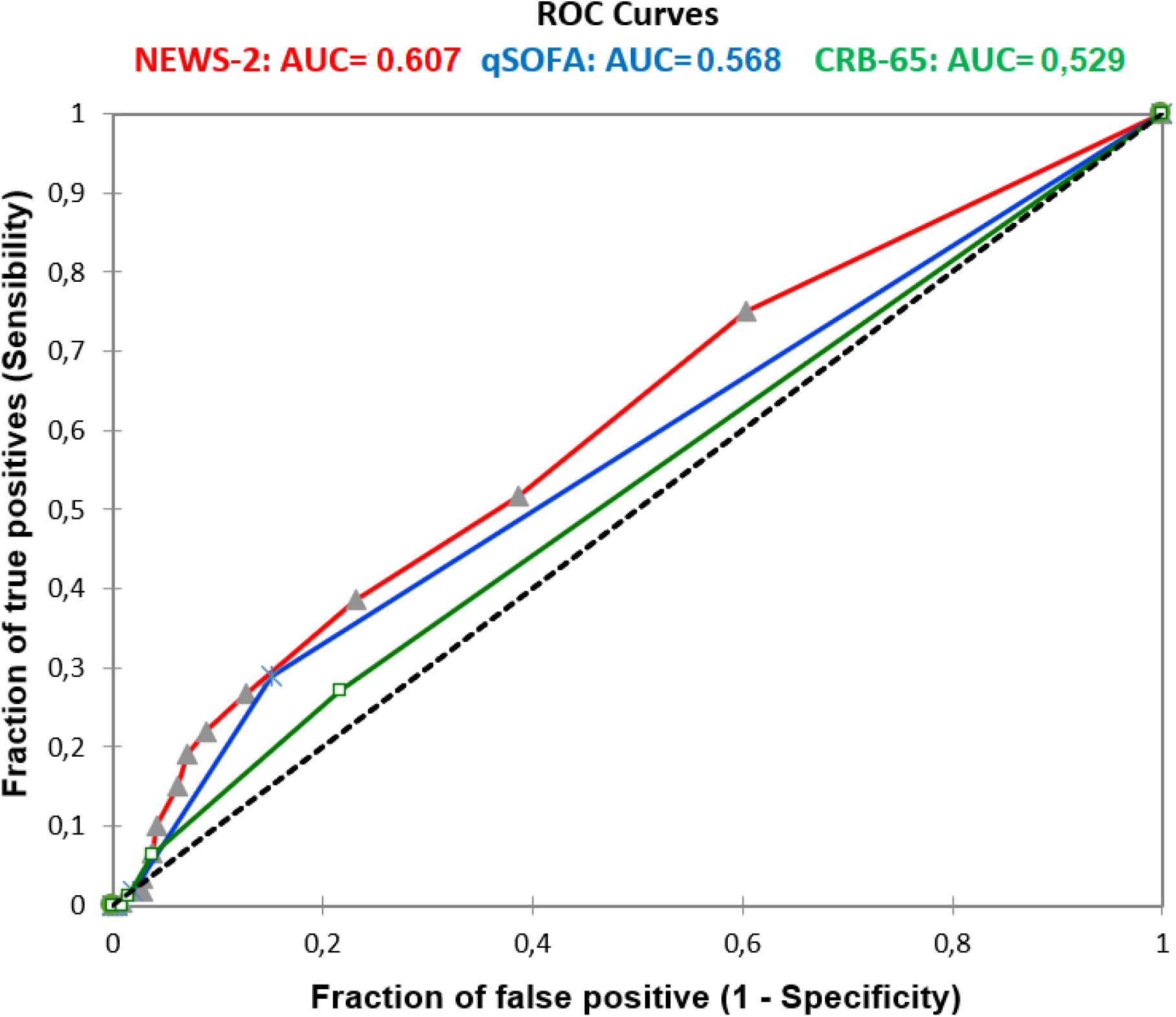
Receiver operating characteristic (ROC) curve of sensitivity vs. specificity for NEWS-2, qSofa and CRB-65 scores for COVID-19 diagnosis.

Of 620 patients for whom we could calculate NEWS-2, 277 underwent a pulmonary CT scan. NEWS-2 reflected the degree of pulmonary involvement (Bravais-Pearson correlation coefficient r = 0.48). NEWS-2 was a good indicator of disease severity in COVID+ patients (Table 6).

**Table 6.** NEWS-2 score means as a function of pulmonary Computed Tomography Scan result (Numbers in parenthesis indicate group sizes; COVID+ = positive with either RT-PCR, or CT scan positive; COVID-= negative with RT-PCR negative, CT scan negative, or negative diagnosis on clinical signs).

|  | NEWS-2 score | NEWS-2 score for COVID-19+ patients | NEWS-2 score for RH patients | NEWS-2 score for HOS patients |
| --- | --- | --- | --- | --- |
| All patients | 2.34 (620) | 2.67 (425) | 1.41 (545) | 7.36 (98) |
| No scan | 1.22 (343) | 1.27 (227) | 1.22 (342) | 3 (1) |
| Scan negative | 2.40 (110) | 2.6 (10) | 1.29 (90) | 7.4 (20) |
| Pulmonary involvement |  |  |  |  |
| Minimum | 3 (38) | 3 (38) | 2.32 (31) | 6 (7) |
| Moderate | 3.64 (45) | 3.64 (45) | 2.49 (35) | 7.70 (10) |
| Spread | 5.24 (41) | 5.24 (41) | 2.33 (18) | 7.52 (23) |
| Severe | 7.03 (32) | 7.03 (32) | 0.5 (2) | 7.47 (30) |
| Critical | 7.80 (5) | 7.80 (5) | 3 (1) | 9 (4) |

If we consider patients with NEWS-2 >5 (optimum cut-off value for disposition), only a few parameters were significantly different between COVID+ and COVID-patients: pulse pressure (p-value =0.02), temperature (*p-value* =0.00016) and degree of pulmonary involvement (p-value <10^−6^).

### COVID diagnosis and disposition

Although HOS patients more often had a confirmed diagnosis than DH patients (78% vs. 60%, respectively), a diagnosis of COVID-19 was not a discriminating factor for disposition.

### Improvement of NEWS-2

The question was how can NEWS-2 be improved in order to use it for either patient disposition or for the diagnosis of COVID-19 severity without losing its simplicity (rapid to perform and calculate, and easy to interpret) and without changing the nature of the score (i.e. a score based on a patient’s vital signs only). Using a cut-off of 5, NEWS-2 is already an excellent predictor of HOS since it concurred with 96% of dispositions made from our ED. However, it is a less good predictor of COVID-19 diagnosis, whatever the cut-off chosen. A Principal Component Analysis (PCA) with the six variables included in NEWS-2 (Table 7) indicates a first axis that explains 33% of the variance and gives the influence of the different components in this variance. All components are standardised for ease of interpretation. O_2_, SpO_2_ and respiratory rate were the most discriminating factors (Table 7).

**Table 7.** Contribution of NEWS-2 score variables to patient orientation.

| Score | Contribution (%) |
| --- | --- |
| O<sub>2</sub> | 31.16 |
| SpO<sub>2</sub> | 30.05 |
| Respiratory rate | 23.10 |
| Mean arterial pressure | 3.66 |
| Pulse rate | 1.71 |
| Temperature | 0.99 |
| Consciousness | 6.32 |

It may be possible to improve NEWS-2 by adding coefficients to the sub-scores, by eliminating non-discriminating components, or by adding new variables.

Multilinear or logistic modelling of ED disposition according to NEWS-2 sub-scores as regressors gave normalised coefficients indicating the major influence of the first three factors (O_2_, SpO_2_, respiratory rate sub-scores) (Table 8).

**Table 8.**
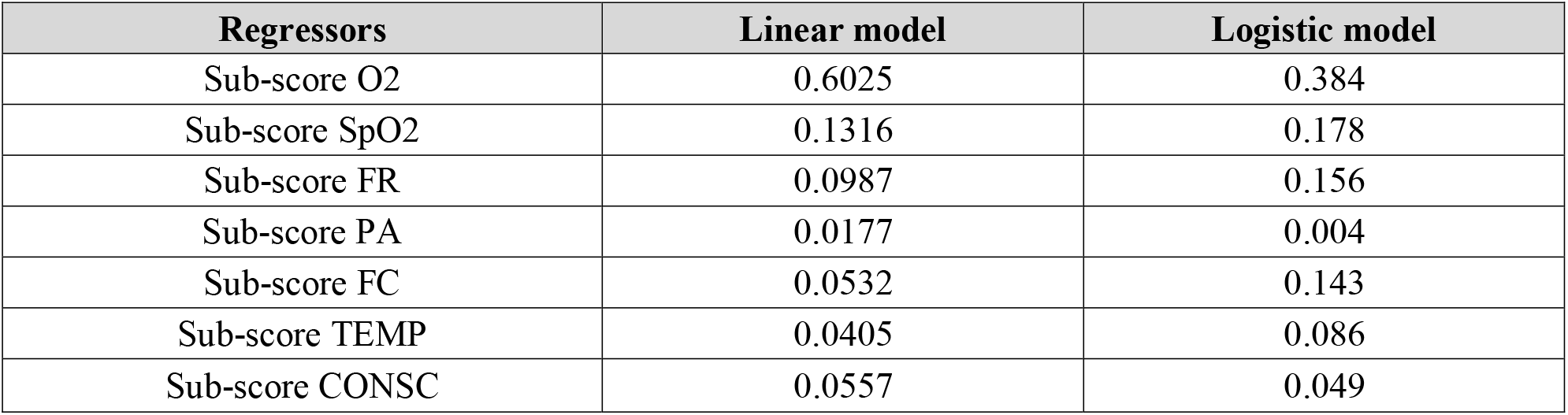
Normalized coefficients for linear and logistic models for NEWS-2 score.

| Regressors | Linear model | Logistic model |
| --- | --- | --- |
| Sub-score O<sub>2</sub> | 0.6025 | 0.384 |
| Sub-score SpO<sub>2</sub> | 0.1316 | 0.178 |
| Sub-score FR | 0.0987 | 0.156 |
| Sub-score PA | 0.0177 | 0.004 |
| Sub-score FC | 0.0532 | 0.143 |
| Sub-score TEMP | 0.0405 | 0.086 |
| Sub-score CONSC | 0.0557 | 0.049 |

The result of the linear model gave a highly discriminatory distribution. With a cut-off of 0.5, the model included 26 patients who were misclassified with respect to real disposition: 9 false positives (DH classified as HOS by regression) and 17 false negatives (HOS classified as DH by regression): compared to the initial NEWS-2 (with a cut-off of 5) the model did not show any significant improvement to predict real disposition from the ED.

Discriminant factor analysis of disposition according to the parameters used in NEWS-2 did not yield a better prediction than NEWS-2 itself, even when age and sex were added (42 misclassified: 30 false negative, 12 false positive) (Table 9).

**Table 9.** Discriminant factor analysis: results.

| from \ to | HOS | RH | Total | % correct |
| --- | --- | --- | --- | --- |
| HOS | 66 | 30 | 96 | 68.75 % |
| RH | 12 | 508 | 520 | 97.69 % |
| Total | 78 | 538 | 616 | 93.18 % |

### Adding new variables

Some potential predictors not present in NEWS-2, including age, comorbidities (e.g. hypertension, coronary disease, stroke, diabetes) were considered in a multilinear regression model to supplement and improve NEWS-2 for the current situation, but the results were not conclusive. As observed, ‘altered condition’ was highly discriminatory, but did not really fit into the philosophy of a score that must be calculated from vital measurements only (recorded by the nursing staff or by a TMB) and not from an assessment of the patient’s condition that requires a physician’s judgment. In addition, biological parameters were not taken into consideration for NEWS-2 improvement as it was considered that the simplicity of NEWS-2 and its independence from laboratories and limited resources had to be preserved.

## Discussion

The main finding of this study was that NEWS-2, used in the ED, was effective at predicting the risk of hospital admission in patients with suspected COVID-19. The study population was selected after an initial triage based on COVID-19 symptoms. NEWS-2 calculated at admission to the ED quickly and easily identified suspected COVID-19 patients at risk of clinical deterioration without medical intervention, proving its usefulness at increasing patient safety, management and monitoring. NEWS-2 >5 predicted hospital admission with 82% sensitivity and 98% specificity. Our data are consistent with recent publications evaluating NEWS-2 in COVID populations that demonstrate its effectiveness at predicting severe disease, in-hospital mortality and admission to an ICU.^24-28^ In our study, NEWS-2 appeared to be superior to the qSOFA and CRB-65 scores at predicting hospital admission of COVID-19 patients, as reported previously.^27,29-32^

Of the 814 patients included in this study, 13.7% were hospitalised, which is lower than the figures found in the literature (20%)^33^ and is one of the major limitations of the study. This can be explained by the fact that the patients self-referred and were generally younger (mean age 46.8-years). Thus, our conclusions on the interpretation of NEWS-2 refer to this population and are not representative of the overall hospitalised population.

Using NEWS-2 >5 as the criterion for disposition misclassified 25 patients. For patients with dyspnoea or acute pain, NEWS-2 was positively affected by pulse rate and respiratory rate. The number of DH patients with a high NEWS-2 could have been reduced by repeating the score after analgesic treatment or reassurance. Elderly patients were more often admitted to hospital despite a low NEWS-2, as their general condition (e.g. anorexia, vomiting, dehydration), degree of pulmonary damage and associated risk factors (e.g. diabetes, high blood pressure, stroke, immunosuppression, myocardial infarction history and chronic lung disease) seem to be important elements for the ED team deciding on hospital admission. Social isolation was another risk factor taken into account by ED practitioners. The addition of these parameters to NEWS-2 could have an influence on decisions for hospital admission, but would add complexity to data recording and processing.

The study population was selected using a clinical questionnaire at the ED entrance to reflect the day-to-day organisation and advanced triage of the emergency services in France during the pandemic. Hospitalising potentially severe COVID-19 cases using a standardised NEWS-2 rather than a COVID biological diagnosis has the advantage of taking into consideration other diseases as well. NEWS-2 could be used as a general tool for admitting seriously ill patients but also for monitoring COVID-19 patients at home. This score has been shown to be effective in both the primary and secondary healthcare settings in several studies.^14, 35-36^

NEWS-2, as its name illustrates, was conceived to detect signs of physiological failure and not as a specific diagnostic tool. Future research might take into consideration the association of NEWS-2 with other scores. To associate a generic score for physiological failure with other tools that are disease-aetiology specific might be extremely helpful. Such a two-step procedure would allow the early detection of patients requiring urgent medical attention while accurately orienting a patient pre-diagnosis.

An interesting perspective is the automatic calculation of NEWS-2 by a TMB integrating autonomous recording of vital signs and the collection of personal history (telemedical record). This allows early self-evaluation of patients and appropriate dispensation. It could be a powerful tool in hospital settings, but even more efficient if deployed at the pre-hospital level as part of a telemedical network. During an epidemic, it would allow massive, rapid, easy and partially health-personnel-free screening and follow-up of the general population (data recording) while reducing patients’ mobility. Minimal risk patients could be excluded from a hospital-based evaluation and high-risk patients could be detected promptly and transferred to the appropriate facilities.^16, 36-41^ This could help regulate the flow of patients to hospitals, predict hospital admission rates and prevent or reduce ED overcrowding. An additional benefit could come from improvements in data collection for epidemiological studies and public health planning.

## Conclusion

NEWS-2 was a useful tool for the evaluation of patients with suspected COVID-19, helping medical practitioners to predict and make decisions on hospital admission. NEWS-2 as a criterion for hospital admission of COVID-19 patients was not improved by the addition of other patient variables, such as comorbidities or age. However, its association with other scores might improve and facilitate intra-hospital evaluation by adding sickness-specificity. The automatic calculation of NEWS-2 by a TMB provided with the appropriate devices and algorithms could be a major improvement in patient management, follow-up and screening, particularly in epidemic situations.

## Data Availability

Database is available on request if confidentiality is ensured.

## Notes

### Competing Interest Statement

The authors have declared no competing interest.

### Funding Statement

No external funding was received

### Author Declarations

The project (referenced under the number COS-RGDS-2021-05-001-FAURE-V) has been submitted to the members of the Scientific Committee of the GCS Ramsay Sante for Education and Research. The committee has reviewed the following documents: application form for ethical advice on a project research and the synopsis of the study. This study has received a favorable opinion on 05/04/2021 (Numero IORG de Ramsay sante Recherche & Enseignement: IORG0009085; Numero IRB du Conseil d'Orientation Scientifique: IRB00010835). All patients were informed of the data collection by a specific poster in the waiting room and gave their informed consent prior to the recording of vital signs by the TMB. All patients received a letter informing them of the study and gave their informed consent, according to MR004 methodology.

